# Gamma Frequency Sensory Stimulation in Probable Mild Alzheimer’s Dementia Patients: Results of a Preliminary Clinical Trial

**DOI:** 10.1101/2021.03.01.21252717

**Authors:** Diane Chan, Ho-Jun Suk, Brennan Jackson, Noah Pollak Milman, Danielle Stark, Elizabeth B. Klerman, Erin Kitchener, Vanesa S. Fernandez Avalos, Arit Banerjee, Sara D. Beach, Joel Blanchard, Colton Stearns, Aaron Boes, Brandt Uitermarkt, Phillip Gander, Matthew Howard, Eliezer J. Sternberg, Alfonso Nieto-Castanon, Sheeba Anteraper, Susan Whitfield-Gabrieli, Emery N. Brown, Edward S. Boyden, Bradford Dickerson, Li-Huei Tsai

**Author notes:** These authors contributed equally (Diane Chan, Ho-Jun Suk, Brennan Jackson, Noah Pollak Milman).

## Abstract

Non-invasive <u>G</u>amma <u>EN</u>trainment <u>U</u>sing <u>S</u>ensory stimuli (GENUS) at 40Hz reduced Alzheimer’s disease (AD) pathology, prevented cerebral atrophy and improved performance during behavioral testing in mouse models of AD. We report data from a safety study (NCT04042922) and a randomized, placebo-controlled trial in participants with probable mild AD dementia after 3 months of one-hour daily 40Hz light and sound GENUS (NCT04055376) to assess safety, compliance, entrainment and possible effects on brain structure, function, sleep and cognitive function. GENUS was well-tolerated and compliance was high in both groups. Electroencephalography recordings show that our GENUS device safely and effectively induced 40Hz entrainment in cognitively normal subjects and participants with mild AD. After 3 months of daily stimulation, participants with mild AD in the 40Hz GENUS group showed less ventricular enlargement and stabilization of the hippocampal size compared to the control group. Functional connectivity increased in both the default mode network and the medial visual network after 3 months of stimulation. Circadian rhythmicity also improved with GENUS. Compared to controls, the active group performed better on the face-name association delayed recall test. These results suggest that 40Hz GENUS can be used safely at home daily and shows favorable outcomes on cognitive function, daily rhythms, and structural and functional MRI biomarkers of AD-related degeneration. These results support further evaluation of GENUS in larger and longer clinical trials to evaluate its potential as a disease modifying therapeutic for Alzheimer’s disease.

## INTRODUCTION

Alzheimer’s disease (AD) is a multi-faceted neurodegenerative disorder characterized by excessive accumulation of amyloid-beta and phosphorylated tau tangles as major pathological features (Canter, Penney, and Tsai 2016). In addition to this molecular pathology, disruptions in neuronal network oscillations are observed in AD (Uhlhaas and Singer 2006; Nimmrich, Draguhn, and Axmacher 2015; Herrmann and Demiralp 2005; Palop and Mucke 2016). For example, gamma band (30-80 Hz) oscillations, which are related to cognitive functions such as attention and memory (J.E. Lisman and Idiart 1995; Engel, Fries, and Singer 2001; John E Lisman 1999) are altered both in human patients with AD (Jing Wang et al. 2017; Van Deursen et al. 2008; Ribary et al. 1991; Jelles et al. 2008; Koenig et al. 2005; Stam et al. 2002) and mouse models of the disease (Verret et al. 2012; Gillespie et al. 2016; Iaccarino et al. 2016). Increasing gamma band oscillations through genetic modifications or optogenetic stimulation can reduce amyloid levels and improve memory in AD model mice (Verret et al. 2012; Martinez-Losa et al. 2018; Etter et al. 2019). However, the relationship between AD pathology and disrupted gamma band oscillations is not yet fully elucidated.

Recently, we discovered that entraining gamma frequency oscillations using light flickering at 40 Hz reduced amyloid load and induced glial response in the visual cortex of AD model mice, effectively attenuating AD-related pathology (Iaccarino et al. 2016). Gamma frequency oscillations were entrained across multiple brain regions beyond the visual cortex and preserved neuronal as well as synaptic densities across the brain areas that showed entrainment; mice also showed and improved cognitive performance (Adaikkan et al. 2019). In another study, we found that auditory stimulation using trains of tones repeating at 40 Hz can induce gamma frequency neural activity and ameliorate pathology and that combined 40 Hz visual and auditory stimulation produces enhanced beneficial effects in AD model mice (Martorell et al. 2019). These studies suggest that entraining gamma frequency oscillations using sensory stimulation (i.e., <u>G</u>amma <u>EN</u>trainment <u>U</u>sing <u>S</u>ensory stimuli or GENUS) can effectively ameliorate pathology and improve cognition in these mouse models and suggests that GENUS is worth pursuing as a therapeutic avenue for the treatment of AD. Given the non-invasive nature of GENUS, we translated this technology for use in humans. We hypothesize that inducing 40Hz entrainment in patients with probable mild AD dementia may modify pathophysiology related to neurodegeneration, leading to improved memory and function.

Previous studies investigating the use of sensory stimulation to induce entrainment showed that 40Hz auditory stimuli created the highest response as measured by electroencephalography (EEG) and increased regional cerebral blood flow in healthy participants (Pastor et al. 2002). Other neuromodulation modalities such as transcranial alternative current (TACS) have also been used to induce 40Hz oscillations to improve abstract reasoning, working memory and insight in cognitively normal people (Emiliano Santarnecchi et al. 2013; E. Santarnecchi et al. 2016; Emiliano Santarnecchi et al. 2019). These results confirm that 40Hz is the optimal frequency to evaluate in humans whether induced gamma entrainment can be applied to slow or prevent neurodegeneration and improve cognition.

We report here two studies of 40Hz combined visual and auditory GENUS: (i) testing target engagement in cognitively normal adults and patients with mild AD dementia (NCT04042922) and (ii) a single-blinded, randomized, placebo-controlled trial testing the safety and efficacy of 40Hz GENUS in patients with mild probable AD dementia (NCT04055376). The main objective of this paper is to report on the feasibility and safety of daily, at-home light and sound GENUS in this population, and target engagement (40Hz entrainment) in cognitively normal adults and patients with AD dementia. The secondary objectives were to explore the effects of daily GENUS on cognition, circadian rhythms, and AD biomarkers including structural and functional MRI.

## RESULTS

GENUS development occurred in two studies: (i) an initial safety and feasibility study to optimize entrainment during stimulation with our light and sound device and (ii) a single-blinded, randomized, sham-controlled trial to evaluate for safety of daily stimulation, compliance of usage and exploratory measures to test if pathophysiology related to neurodegeneration could be modified using 40Hz GENUS (demographics described in Table 1 and 2).

**Table 1.**
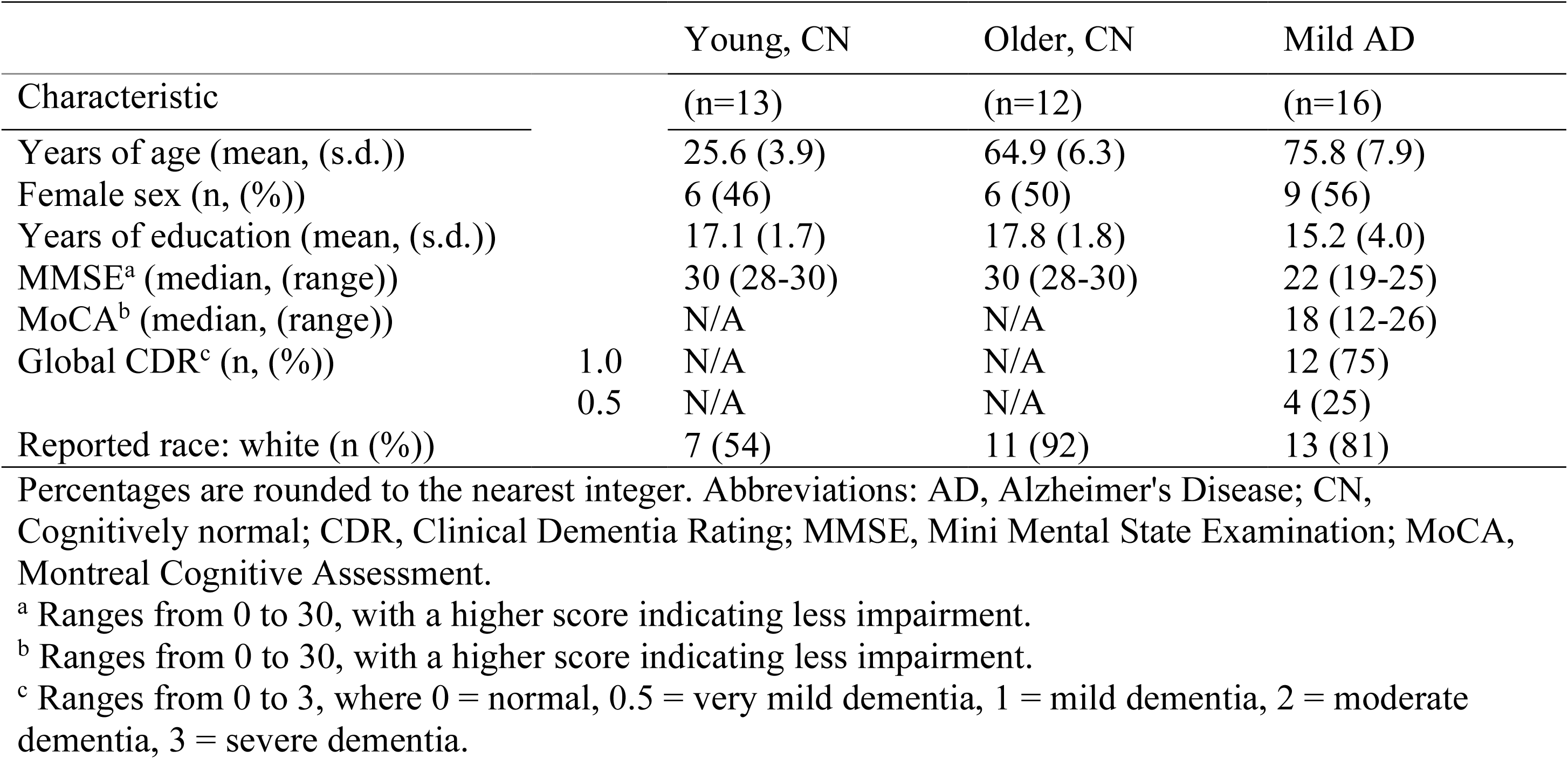
Demographics and Baseline Clinical Characteristics of Acute Stimulation Participants.

**Table 2.** Adverse Events of Acute and Longitudinal GENUS Stimulation in Participants

| Adverse Events | Acute Stimulation |  |  | Longitudinal Stimulation |  |
| --- | --- | --- | --- | --- | --- |
|  | Young, CN<br>(n= 13) | Older, CN<sup>a</sup><br>(n= 10) | Mild AD<br>(n= 16) | Mild AD<br>Control (n= 7) | Active (n= 8) |
| New Onset Headache (n, (%)) | 0 | 1 (10) | 0 | 0 | 0 |
| Dry Eye (n, (%)) | 1 (8) | 0 | 0 | 0 | 2 (25) |
| Light Sensitivity (n, (%)) | 1 (8) | 1 (10) | 1 (6) | 0 | 1 (13) |
| Nervousness or anxiety (n, (%)) | 0 | 0 | 0 | 3 (43) | 1 (13) |
| Sleepy or Drowsy (n, (%)) | 5 (38) | 4 (40) | 1 (6) | 1 (14) | 3 (38) |
| Numb / Unfocused (n, (%)) | 0 | 0 | 0 | 1 (14) | 2 (25) |
| Bored (n, (%)) | 0 | 0 | 0 | 0 | 1 (13) |
| Nausea (n, (%)) | 1 (8) | 0 | 0 | 0 | 0 |
| Seizures (n, (%)) | 0 | 0 | 0 | 0 | 0 |
| > 1 Adverse Events (n, (%)) | 1 (8) | 1 (10) | 0 | 2 (29) | 4 (50) |
| No Adverse Events (n, (%)) | 6 (46) | 5 (50) | 14 (88) | 3 (43) | 3 (38) |

Percentages are rounded to the nearest integer. Abbreviations: AD, Alzheimer's Disease; CN, Cognitively normal.
<sup>a</sup> Two Older, CN participants were excluded from this analysis due to COVID-19 Shutdown.

### Optimization of GENUS for safety and induced entrainment

To assess entrainment of neural oscillations by 40 Hz visual, auditory or synchronized visual and auditory stimulation and to assess for safety, we exposed young, cognitively normal adults (age 18-35), older cognitively normal adults (age 50-100) and patients with mild AD dementia to 40Hz GENUS while recording their scalp EEG (Table 1, Fig. 1). The 40Hz GENUS device is comprised of a 2 feet x 2 feet light panel and speaker that is programmable to deliver light or sound alone at 40Hz, synchronized light and sound at 40Hz and control settings with constant light with or without white noise. Testing was done with a small tablet at the center of the light panel showing videos for entertainment and to engage attention to the center of the light source. In all groups, GENUS with synchronized light and sound at 40Hz significantly increased the 40 Hz power spectral density (PSD) from the mock-stimulation level (constant light and white noise) at both the frontal and the occipital electrode sites, engaging brain regions not responding to visual or auditory stimulation alone (Figure 1A and 1B). The median change in the 40 Hz PSD at the frontal electrode site was 7.71 dB (range, 0.68 to 16.88; p < 0.001) for the cognitively normal young group, 7.22 dB (range, 0.79 to 13.10; p = 0.002) for the cognitively normal older group, and 5.82 dB (range, -0.02 to 10.51; p < 0.001) for the mild AD group; the median change in the 40 Hz PSD at the occipital site was 7.74 dB (range, 3.51 to 19.45; p < 0.001) for the cognitively normal young group, 7.95 dB (range, 1.18 to 14.70; p = 0.002) for the cognitively normal older group, and 7.68 dB (range, 4.23 to 18.26; p < 0.001) for the mild AD group. Harmonics at 80Hz and sub-harmonics at 20Hz can be seen in the cognitively normal younger and older groups, and less so in the mild AD group, which is consistent with what is reported in the literature (Kikuchi 2002). The increases in 40 Hz PSD were accompanied by significant increases in 40 Hz coherence (Figure S1A, B, C and Table S2), indicating that the combined stimulation induced synchronized 40 Hz neural oscillations across multiple electrode sites in cognitively normal participants and patients with mild AD dementia.

**Figure 1.**
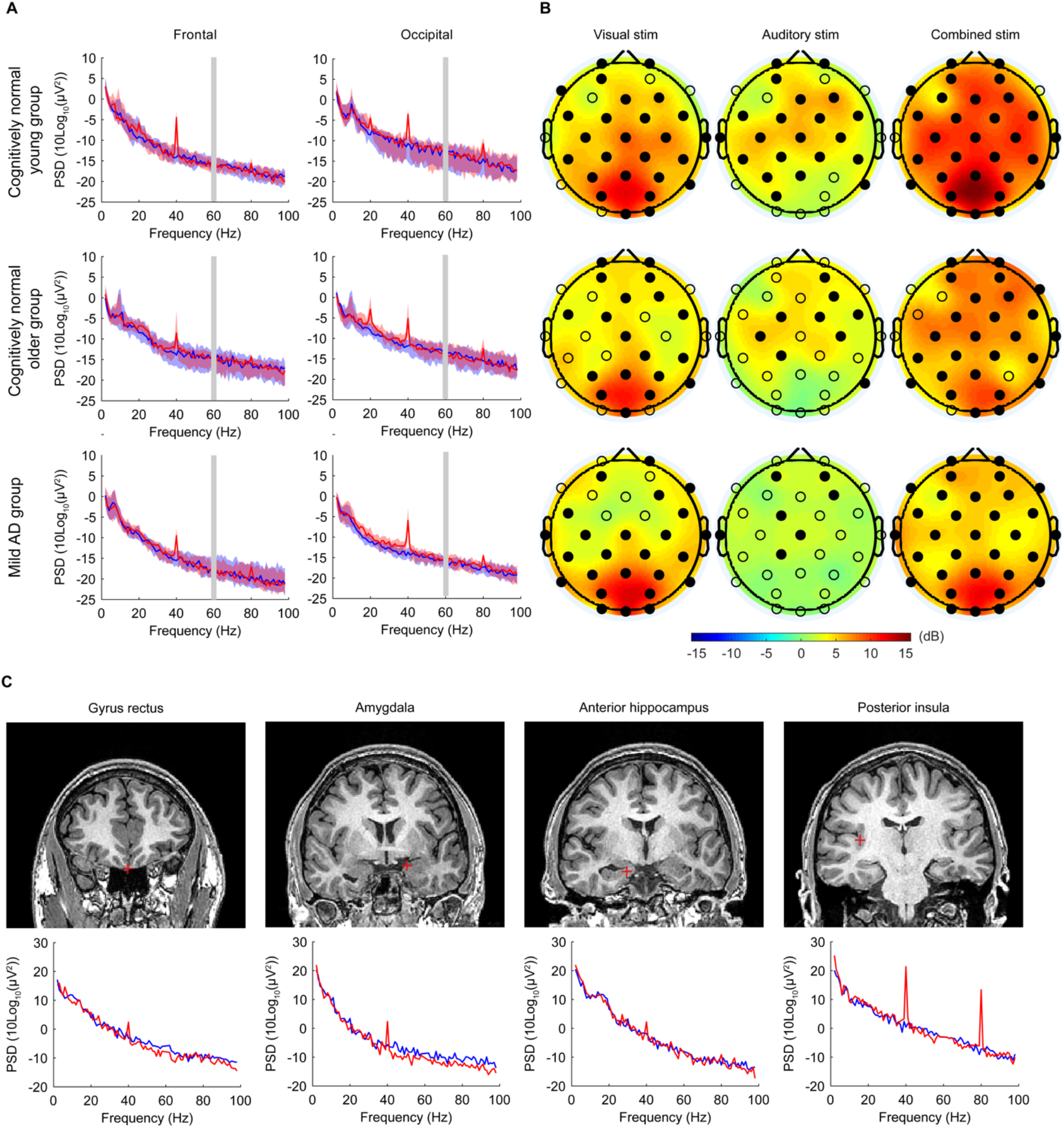
Acute 40 Hz Combined Visual and Auditory Stimulation Entrains Cortical and Subcortical Regions. (A) Scalp EEG power spectral density (PSD) at the frontal (Fz, F3, F4, F7, F8) and the occipital (Oz, O1, O2) electrode sites, in cognitively normal young subjects (n = 13; top row), cognitively normal older subjects (n = 12; middle row), and patients with mild AD (n = 16; bottom row). Solid lines, group median; shaded areas, 95% confidence interval; blue, no stimulation; red, 40 Hz combined stimulation. Gray bar placed around frequency range with 60 Hz line noise. (B) Topographic maps showing the median change in 40 Hz PSD from the no-stimulation level with 40 Hz visual alone, 40 Hz auditory alone, and 40 Hz combined stimulation, in cognitively normal young subjects (n = 13; top row), cognitively normal older subjects (n = 12; middle row), and patients with mild AD (n = 16; bottom row). Filled circles represent scalp electrodes at which the increase in 40 Hz PSD from the no-stimulation level was significant (p < 0.05, Wilcoxon’s sign rank test, Bonferroni corrected for 32 electrodes). (C) Example coronal MRI images before electrode implantation (top row) and intracranial EEG power spectral density (PSD; bottom row) from a single patient with medically intractable epilepsy (Patient 483), for depth electrode contacts placed in the gyrus rectus, amygdala, anterior hippocampus, and posterior insula. Red plus sign, approximate location of the depth electrode contact; blue, no stimulation; red, 40 Hz combined stimulation. For the PSD between 58 Hz and 62 Hz, interpolated values are plotted, because bandstop filtering around 60 Hz during signal preprocessing (for line noise removal) led to a large dip in the PSD around 60 Hz.

In addition to our trials, we also assessed the effects of 40Hz GENUS light and sound in two patients with epilepsy who had implanted intracranial EEG leads for epilepsy surgical planning purposes to evaluate for target engagement within the brain during stimulation. Intracranial EEG shows that 40 Hz light and sound GENUS entrains deeper regions of the brain, including the gyrus rectus, amygdala, hippocampus, insula and in more superficial areas such as frontal and temporal gyri (Fig. 1C and Figure S2A, B).

The largest increases in 40 Hz PSD were detected in the insula and the superior temporal cortex, which have been shown to be involved in multi-sensory integration (Bushara, Grafman, and Hallett 2001; Calvert 2001). Across subcortical and deep cortical regions, there was an overall increase in 40 Hz PSD during the combined stimulation compared to the no-stimulation period (median [range] change in the 40 Hz PSD from the no-stimulation level: 4.58 dB [-1.22 to 23.27] in Patient 483; 0.77 dB [-1.95 to 6.44] in Patient 493), with the increase in 40 Hz PSD appearing at the majority of the electrode contacts (Figure S2A; 75/78 contacts in Patient 483; 10/15 contacts in Patient 493). Additionally, the combined stimulation led to an increase in 40 Hz coherence not only between nearby electrode contacts but also between deep and superficial electrode contacts (Figure S2C, D), suggesting that the stimulation led to synchronized 40 Hz oscillations across deep and superficial brain areas, including those beyond primary sensory areas. GENUS was well tolerated by all subjects with no significant adverse events as evaluated by adverse events questionnaire and by EEG analysis performed by an independent epileptologist, which showed no seizure activity (Table 2).

### Longitudinal Study Design

We conducted a longitudinal, randomized, placebo-controlled study to evaluate long-term effects of GENUS for safety and compliance in participants with probable mild AD dementia. Participants with mild probable AD dementia were recruited and randomized into control and active arms (Table 3; n = 7, n = 8, respectively; see Methods for full details). Both groups were given the same light and sound devices described above with a programmable light panel and speaker delivering the same light and sound level, with a tablet for entertainment. The control device delivered constant white light and white noise while the active device delivered 40Hz synchronized light and sound. The light and sound level delivered by both the control and active settings were the same. The device was used by participants at home for 1 hour daily. Compliance was measured using a built-in timestamp indicating when the device was on and through a mounted camera, which took pictures of the participant every 5 seconds when the device was turned on. Participants were evaluated with EEG for entrainment, both structural and functional magnetic resonance imaging (MRI) of the brain, and cognitive assessments at baseline and after 3 months of at-home device usage. Actigraphy watches worn by participants constantly through the study period recorded sleep and activity. Safety was assessed with EEG at baseline and 3 months and weekly phone calls to assess safety with an adverse events questionnaire. The participants continued to use the GENUS devices at home daily and compliance, actigraphy and safety was assessed as described until the end of the planned trial at 9 months. The study had another planned clinical assessment (EEG, MRI, cognitive battery) at 9 months, but due to the COVID-19 pandemic, this visit was not possible. Cognitive assessment was done virtually at 9 months but not reported here, since the testing modality was different than that used in the earlier time points.

**Table 3.**
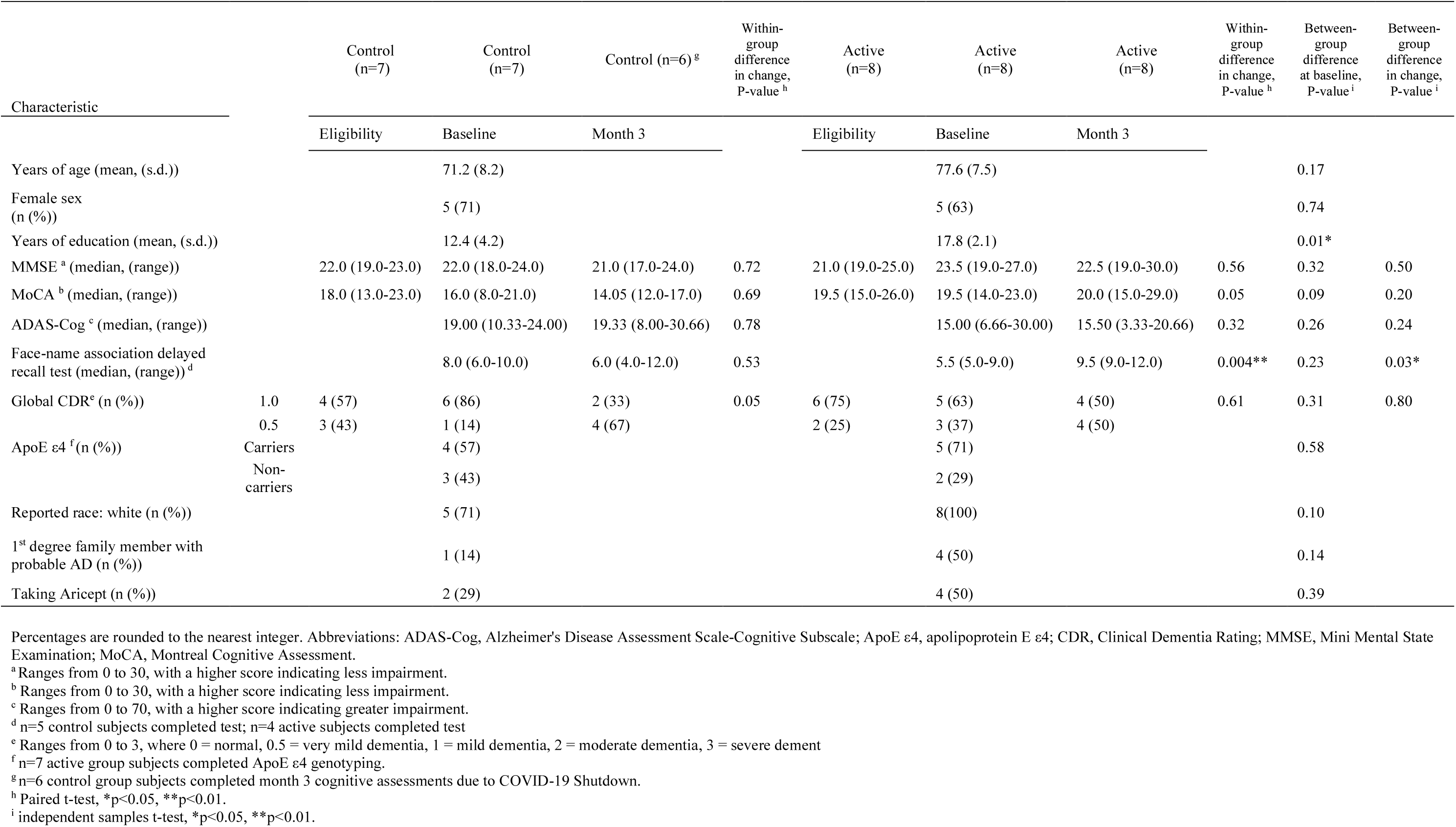
Demographics and Clinical Characteristics of Longitudinal Stimulation Participants Diagnosed with Probable Mild Alzheimer’s Disease.

A total of 87 potential participants were screened, of whom 56 were excluded as they did not meet study criteria, 12 declined to participate or did not respond and 4 did not participate for other reasons (see Methods for inclusion/exclusion criteria, Table S1). Fifteen participants were enrolled in the longitudinal study, of whom 8 completed the active arm and 7 completed the control arm. A CONSORT diagram of the study flow can be found in (Figure S3). Patient characteristics at baseline, including age, sex, APOE status, and cognitive scores were not statistically different across the two trial groups, with the exception of years of education, in which the control group had significantly fewer years of education compared to the active group due to a single outlier (Fig. S4, Table 3; p = 0.01). However, education was not significantly correlated with any outcome variables of interest, reducing concern that differences in education might be a confounding factor (Fig. S5, S6).

### Safety and Compliance of Usage

Safety was assessed with EEG during stimulation with the GENUS device at baseline and after 3 months to monitor for epileptiform discharges and through weekly phone calls with participants. 40Hz GENUS was well-tolerated by all participants with no significant adverse events (Table 2). Timestamp data and photographic records of participants during device usage acquired as described above were analyzed for compliance. After 4 months of daily stimulation, there was no significant difference in compliance of usage between the groups – mean usage was 91.04% ± 6.84% and 86.81% ± 9.43% for the control and active groups, respectively. Both the control and active groups used their devices at home for an hour daily equally (p = 0.355).

### The GENUS group showed less brain atrophy and increased functional connectivity at 3 months

Cerebral atrophy occurs in the natural progression of AD and ventricular enlargement can be seen in the course of this atrophy. Using structural MRI, we found a significant difference in ventricular enlargement between the control and active group following 3 months of GENUS (p = 0.024) (Fig. 2A and 2B, Table S5). Analysis of change in total ventricular size from baseline to month 3 indicates that the control group exhibited ventricular enlargement (4.34+/-1.72%, p = 0.0016). In contrast, the active group did not have a significant change in ventricular volume (1.33+/-2.33%). The mean Cohen’s d effect size for ventricular enlargement was 0.59, taking into account population variability. Hippocampal (HPC) volume declined in the control group but did not decline in the active group (Fig. 2C; Table S5; bilateral HPC volume (n = 13, control p = 0.034, active p = 0.438). Subjects who did not complete a structural MRI at both baseline and 3-months were not included in this analysis.

**Figure 2.**
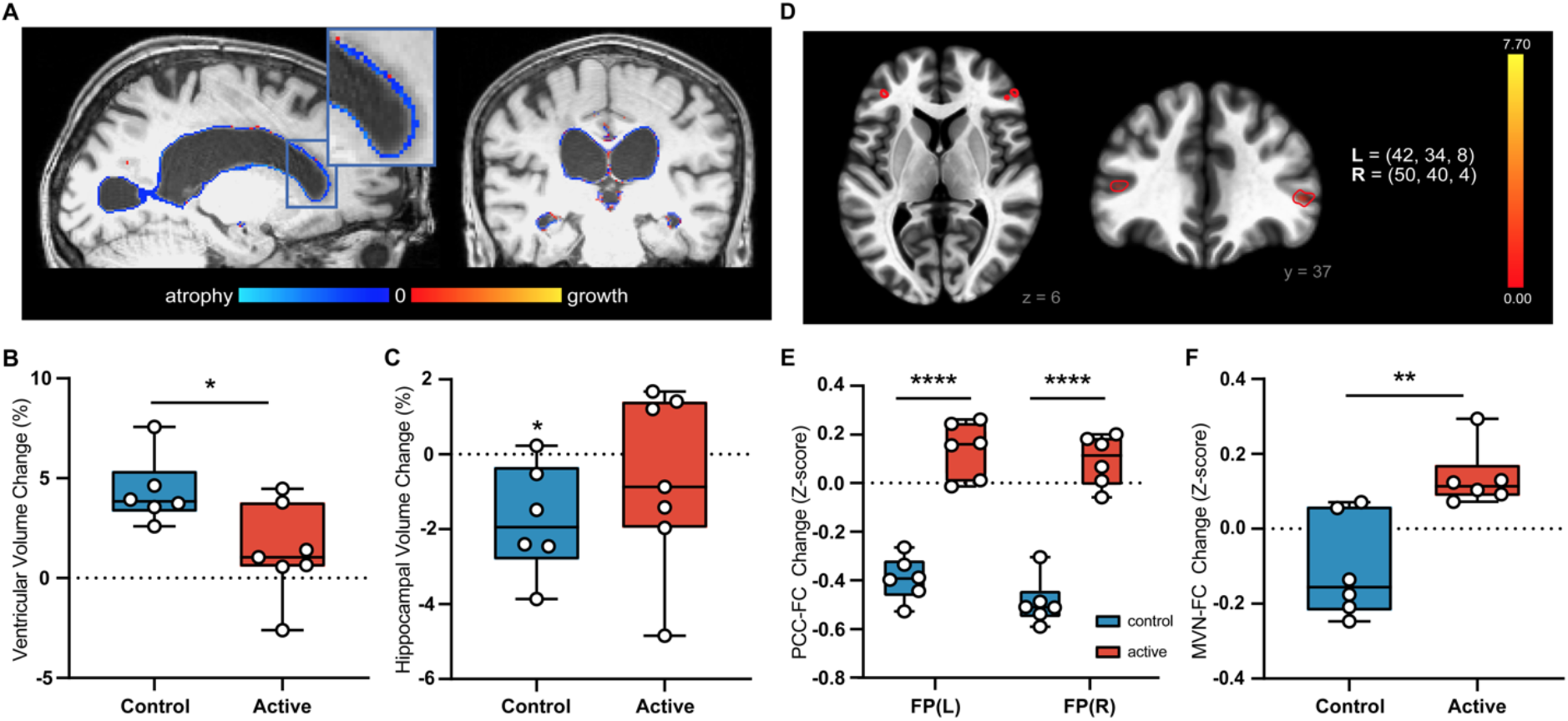
Chronic GENUS leads to group-level differences in structural and functional MRI outcomes. (A) Example image illustrating ventricular enlargement in a control subject from baseline to 3 months. (B) Group-level analysis showing greater ventricular enlargement in the control group compared to the active group (n = 13, p = 0.024, unpaired t-test) (C) Group-level analysis of control vs active HPC volume compared to 0 (n = 13, control p = 0.034, active p = 0.438) (D) Seed-to-Voxel analysis of PCC-FC in control group from baseline to 3-months (See Table S3, p < 0.05 FWE-corrected) (E) Seed-to-voxel analysis of PCC-FC for between group comparisons from baseline to 3-months (n = 12, p < 0.05 FWE-corrected) (F) Group-level analysis of changes in mean functional connectivity of the MVN from baseline to 3-months. (n = 12, p = 0.004, paired t-test) FP: Frontal Pole MVN: Medial Visual Network PCC-FC: Posterior Cingulate Cortex functional connectivity PVVC: Percent Ventricular Volume Change

To evaluate whether neural networks were altered by 40Hz GENUS, we used functional MRI to probe circuits important for memory and sensory processing, including the default mode network (DMN) and medial visual network (MVN), respectively. Subjects were removed from analysis if the number of valid scans made up less than 10% of the total recorded scans due to motion or other artifacts. A seed-to-voxel analysis of the posterior hub of the DMN containing the posterior cingulate cortex (PCC) and the precuneus was performed and differences between baseline and follow-up scans were compared between groups. Between group comparison showed a significant group difference such that the control group (n=6) declined in functional connectivity between the posterior hub of the DMN and bilateral frontal poles while the active group (n=6) did not (paired t-test; Fig. 2E, Table S3). The control group also observed significant loss in functional connectivity between the PCC and the posterior right supramarginal gyrus, left angular gyrus, inferior right frontal gyrus, and superior right frontal gyrus while the active group did not (paired t-test; Table S3).

Seed-to-voxel analysis of the medial visual network indicated that the active group showed a significant increase in mean functional connectivity of the MVN after three months of GENUS (Fig. 2F; 0.14+/-0.07, p = 0.009). The control group did not show a significant change in mean functional connectivity (−0.11+/-0.14). No difference was observed in baseline values (Table S4). When the functional connectivity was analyzed with the hippocampus as the seed region, the active group had a significant increase in functional connectivity between the left hippocampus and the visual cortex while the control group did not (Table S3, P<0.05, family-wise error (FWE)-corrected).

### The GENUS group showed improved circadian rhythmicity

Prospective studies of circadian rhythms demonstrate reduced stability and increased fragmentation with age, and these changes accelerate with diagnosis of MCI or AD and hasten the transition between the two (Li et al. 2020). Considering its relevance to pathological aging and quality of life, we quantified sleep to better appreciate how a non-invasive light and sound therapy may impact functional outcomes in the progression of AD. We used standardized measurements of stability to characterize day-to-day consistency of activity rhythms, which reflect an individual’s bedtime schedule or ability to become active at a regular time, is a sign of robust coupling to environmental cues. Fragmentation describes sleep/wake transition within a day, characterized by nighttime disturbances or daytime naps which may impact an individual’s subjective sleepiness or ability to complete daily tasks.

At baseline we verified there were no differences between active and control groups in two non-parametric metrics used to quantify circadian rhythmicity using Interdaily Stability (IS; active group mean 0.52, control group mean 0.63, p = 0.83) and fragmentation using Intradaily Variability (IV; active group mean 1.13, control group mean 0.86, p = 0.4) IS and IV are fixed values ranging between 0-1 and 0-2, respectively. For analyses, we used the linear subtraction of each timepoint value minus baseline for each subject. The active group had improved IS over the first four months and significantly improved IS compared to control group after 4 months (active mean 0.072, control mean -0.055, p = 0.036, interaction p = 0.025 with Sidak’s multiple comparisons at M3 p = 0.051 and M4 p = 0.0065) (Figure 3B). The IV of the active group does not change from baseline, while the IV of the control group worsens, although not significantly (active mean -0.029, control mean +0.10, p = 0.22) (Fig. S7).

**Figure 3.**
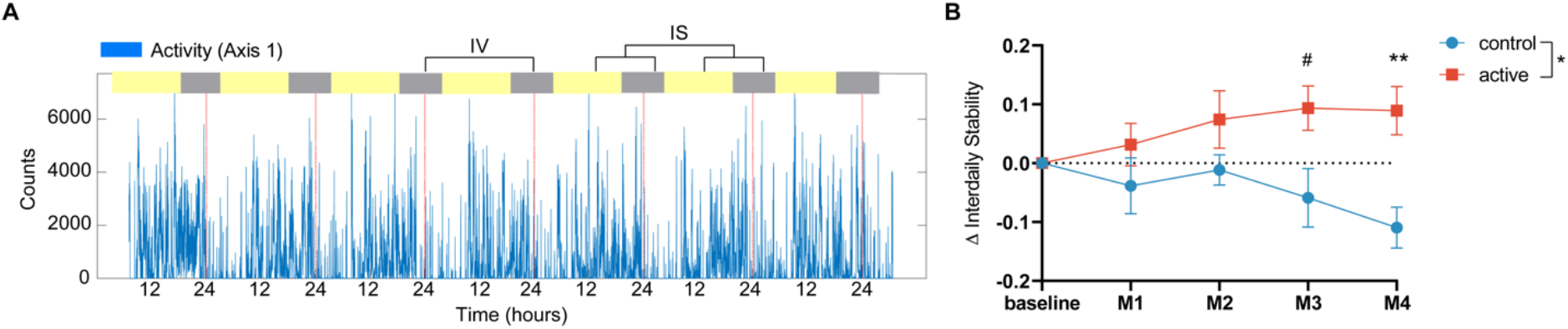
Changes in Rest-Activity Patterns compared to Baseline. (A) Example 7-day activity recording of activity counts (blue) with dotted lines every 24 hours (red). Interdaily Stability (IS) represents the regularity of day-to-day rest-activity patterns. (B) Improved Interdaily Stability compared to baseline in active group but not control group and significant differences between active and control groups at Month 4 and a trend at Month 3. Statistical analysis with a 2-Way ANOVA with Šídák’s multiple comparisons at each monthly timepoint. Lines indicate means ± SEM. Mean of control = -0.055, mean of active = 0.072. #p < 0.1, **p < 0.01. Main effect of intervention p = 0.037, interaction of intervention x time, p = 0.025. Multiple comparison at M3, p = 0.051, M4, p = 0.007.

### The GENUS group showed improved associative memory at 3 months

Cognitive function was assessed at baseline and again during the three-month visit. One participant who was not able to attend both test sessions due to the COVID-19 pandemic was excluded from the analysis, leaving a total of 14 participants (active, n=8; control, n=6). After 3 months of daily 40Hz GENUS, there were no significant differences between groups in cognitive functioning as assessed using the MMSE (p = 0.536), MoCA (p = 0.198), ADAS-Cog (p = 0.237) or Global CDR (p = 0.792), which is not unexpected in this mild AD cohort over a short time frame.

Associative memory tasks have been found to be especially sensitive in the early stages of AD (Parra et al. 2010; Clare et al. 2002; Werheid and Clare 2007; Blackwell et al. 2004). No significant changes in connectivity during encoding of the associative memory task were observed between baseline and after 3 months of stimulation in the fMRI portion of this protocol for either group. At the 3-month visit, the active group (n=4) had a significant improvement in the face-name association recall task performed outside of the MRI (Fig. 4A; 3.75+/-0.96 pts, p = 0.004) while no significant change was observed in the control group (n=5) and there was a significant difference in the change between the control and active groups (p = 0.027). Of interest, the improvement in face-name association recall was significantly correlated with the change in mean functional connectivity of the medial visual network (Fig. 4B; R^2 = 0.52, p = 0.028).

**Figure 4.**
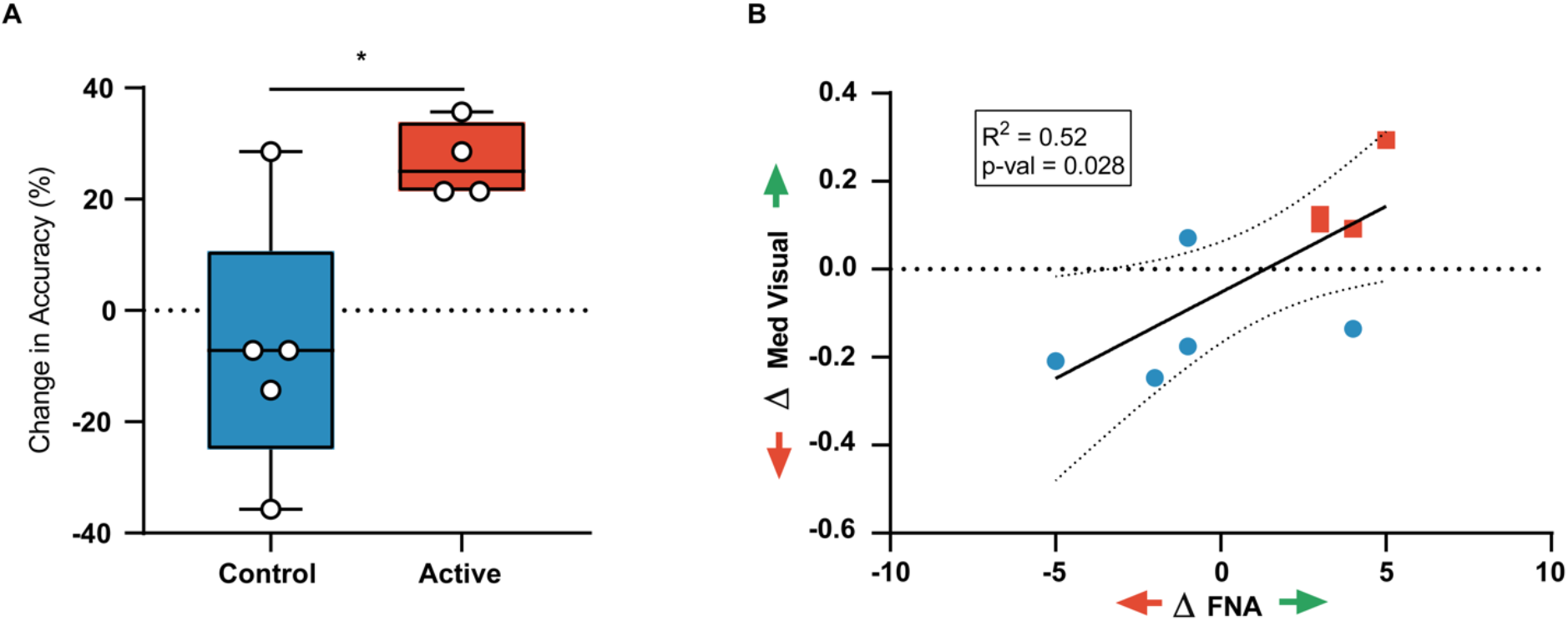
Chronic GENUS improves face-name recall. (A) Group-level analysis of changes in face-name recall test from baseline to 3-months (B) Correlation of change in face-name recall test score with change in mean functional connectivity of the MVN (MVN: Medial Visual Network)

There was a significant difference in years of education between the control and active groups, driven by an outlier who had only 7 years of education in the control group (Table 3, Fig. S4). However, regression analysis relating years of education to outcomes in the face-name association recall task show no significant correlations (Fig. S5A; R^2 0.0003, p = 0.96). Years of education did not correlate with any outcome measures reported including connectivity with MVN (R^2 = 0.097, p = 0.33), change in hippocampal volume (R^2 = 0.16, p = 0.18), change in connectivity with PCC (R^2 = 0.007, p = 0.79) and change in PVV (R^2 = 0.19, p = 0.15) (Fig. S5B, C, D, E).

## DISCUSSION

In AD mouse models, inducing gamma entrainment using 40 Hz sensory stimulation ameliorates pathology and improves cognition (Iaccarino et al. 2016; Adaikkan et al. 2019; Martorell et al. 2019). In our optimization study, we demonstrated that 40Hz GENUS using synced light and sound can effectively induce gamma entrainment across multiple brain regions in cognitively normal individuals, patients with medically intractable epilepsy, and in patients with mild Alzheimer’s disease dementia. We also showed that the combined light and sound stimulation entrains both cortical and subcortical regions, including those that visual or auditory stimulation alone cannot entrain. 40Hz GENUS was safe, does not trigger epileptiform activity even in patients with epilepsy, and does not cause any severe adverse events. In a single-blinded, placebo-controlled RCT in patients with mild AD dementia, we found that relative to the control group, the group that had daily usage of 40Hz GENUS over 3 months had less brain atrophy and reduced loss of functional connectivity, improved markers of circadian rhythmicity and improved performance in an associative memory task. The GENUS device was easy to use for at-home daily stimulation with high compliance in both the placebo and active 40Hz groups and was safe with no significant adverse events. While this preliminary study was powered to evaluate for safety and compliance of usage, the above exploratory outcome measures revealed significant results that were encouraging. Overall, these findings suggest that 40Hz GENUS should be studied more extensively in larger studies to evaluate its potential as a possible disease-modifying intervention in AD.

Previously, we showed that chronic GENUS significantly reduced the expansion of ventricular size and ameliorated neuronal density loss in AD animal models (Adaikkan et al. 2019). In this study, we observed that after 3 months of daily stimulation, the active group showed a lesser degree of ventricular expansion and hippocampal atrophy than the control group. The degree of ventricular expansion and hippocampal atrophy experienced by the control group in our study is consistent with previous studies of mild AD dementia (Ledig et al. 2018). Given that ventricular expansion and hippocampal atrophy correlate with cognitive function and clinical disease progression, this may indicate a slowing or delay in normal progression of the disease (Jack et al. 2009; Nestor et al. 2008; Ledig et al. 2018). However, given the well-established heterogeneity of rates of progression in AD dementia, it is also possible that the two groups would have shown these differences without the intervention. A within-participant cross-over design or a larger parallel-group design would be a logical next step to determine whether these results are robust and reproducible.

Resting-state fMRI has been used to examine functional brain network disruptions in AD that correlate with cognition (Binnewijzend et al. 2012; L. Wang et al. 2006). The network that shows the greatest amount of dysfunction in AD is the default mode network (DMN), which exhibits hypoconnectivity in AD, mild cognitive impairment (MCI), and even those at high risk for AD, especially within its posterior hub – the PCC (Badhwar et al. 2017; Sorg et al. 2007; H. Zhang et al. 2009). This region has been shown to play a critical role in attention, internally-directed thought, and is connected with the medial temporal lobe system and correlated with memory encoding and recall (Leech and Sharp 2014; L. Wang et al. 2010; Miller et al. 2008). Consistent with changes seen in the DMN in the natural progression of AD (Hafkemeijer et al. 2017; H.-Y. Zhang et al. 2010; Jinhui Wang et al. 2013), our control group had significant loss of connectivity between the PCC and several regions in the frontal cortex, as well as the angular gyrus. While declines over such short durations have been previously reported, longer term follow up should provide further insight into the functional changes in the network. Additionally, different neurological disorders besides AD may affect DMN connectivity so larger clinical trials may be needed to confirm these initial results.

The functional connectivity of the visual network was quantified as an area of interest given our mode of intervention, even though it is not typically evaluated in resting-state fMRI studies in AD research. Our work in animal models showed the impact of GENUS on the oscillatory activity and amyloid deposition within the related sensory cortices (Iaccarino et al. 2016; Martorell et al. 2019). Changes in functional connectivity in regions of the brain in AD patients have been shown to be heavily linked with amyloid deposition measured using PET biomarkers and cortical atrophy (Hampton et al. 2020; Myers et al. 2014). A significant increase in mean functional connectivity in the medial visual network following chronic GENUS may indicate an impact on AD pathology within the region or may simply be a result of increased activity from daily use of an intervention though we controlled for this in our placebo stimulation group who used the same intensity and duration of light daily. Very similar face-name association tasks have shown that successful recall of face-name pairs is heavily correlated with amyloid deposition (Rentz et al. 2011). While it was not unexpected that performance on cognitive tasks such as the MOCA, MMSE, ADAS-Cog did not show significant changes over this short time period of 3 months, the improvement in the face-name association task hypothetically may indicate that visually evoked memory may selectively improve due to 40Hz GENUS using visual and auditory modalities but this requires further evaluation. Alternatively, this could be due to the increased sensitivity of the face-name association task in this mild Alzheimer’s dementia group to detect changes in such a short time frame as other studies have shown that this task is a sensitive marker of early amyloid-related impairment (Rentz et al. 2011). A significant increase in the connectivity between the hippocampus and the visual cortex was observed in the active group but not in the control group, which is an interesting avenue to evaluate in future studies as a possible mechanism underlying the improvement observed in this visual-based cognitive task.

Critical biochemical and behavioral changes appear to work bi-directionally between sleep and AD, highlighting the utility of an intervention that impacts sleep (Ju, Lucey, and Holtzman 2014). Sleep is also a crucial mediator of waste clearance including Aβ peptides, facilitated by glymphatic flow and it is hypothesized that changes to sleep architecture, increased nighttime disturbances and reduced REM sleep underlie the accumulation of harmful peptides in both prodromal and AD patients (Nedergaard and Goldman 2020). Because of the nature of our study, we used wrist actigraphy to assess the extent to which GENUS could impact 24-hour activity (Camargos, Louzada, and Nóbrega 2013). GENUS significantly increased day-to-day regularity of activity patterns (as quantified using Interdaily Stability, IS) over four months. Our finding resonates with results reported in the literature suggesting that 40 Hz stimulation can intervene with circadian rhythms that ordinarily degrade with age and more rapidly in AD and can serve as a readout in future GENUS studies (Figueiro and Leggett 2021). Further work on GENUS should include formal studies of sleep using polysomnography in addition to exploring the relationship between improved sleep and clearance of AD pathology.

Little is known about the mechanisms relating gamma stimulation to circadian rhythms. One month of daily hour-long 40 Hz visual stimulation rescues central clock gene expression deficits present in AD mouse models, including CLOCK, BMAL1, and PER2, suggesting 40 Hz stimulation could directly impact the suprachiasmatic nucleus (SCN) (the site of the circadian pacemaker in mammals) and underlying circadian rhythms (Yao et al. 2020). These genes are critical regulators of the circadian system and are posited to explain age and disease related patterns in activity levels. Age and AD-related physiological changes impair nominal biological clock activity and emphasize the overlap of sleep disturbances, behavioral disturbances and quality of life (Figueiro 2017). Improving individual’s rest-activity patterns with 40Hz stimulation and in turn, maintaining consolidated sleep may promote resilience to disease progression. Employing wearable devices to track changes in rest-activity patterns like Interdaily Stability is appropriate for this study population, although more robust measures of sleep may be required to fully appreciate the effects of 40Hz. In examining participant’s specific chronotypes, there is opportunity to personalize the timing of stimulation paradigms to improve the effects on circadian rhythms.

Manipulation of neural oscillations to augment gamma entrainment using 40Hz GENUS was shown to broadly and robustly reduce AD pathology and improve cognition in AD mouse models. Our study is the first to show that 40Hz GENUS induced gamma entrainment can be done in patients with mild AD dementia, and may be associated with positive cognitive and biomarker outcomes, while recognizing the limitations of interpreting results from such a small sample. Our analyses used robust methodology to control for multiple comparisons and important cofounders. However, we must acknowledge some limitations to our study including our small sample size, which made it difficult to conclusively delineate contributions of ApoE4 on functional connectivity and sleep outcomes. In addition, while our intervention was continued at home for a full 9 months as planned, the COVID-19 pandemic interfered with data collection for MRI and EEG after 3 months of stimulation. Despite these limitations, our study supports the possibility that 40Hz GENUS should be studied in larger, longer RCTs in AD, including future studies investigating effects of 40Hz GENUS on amyloid and tau biomarkers of AD.

## Data Availability

All data are available in the main text or the supplementary materials.

## Acknowledgments

We thank Emily Niederst for her careful reading, insight and comments on the paper.

## Author contributions

Conceptualization: LHT, DC, HJS, BD, ENB, ESB

Methodology: DC, HJS, BJ, EBK, NPM, DS, SDB, SWG

Investigation: DC, HJS, BJ, NPM, DS, SDB, EK, VSFA, AB, BU, PG, MH, AB, EJS, ANC, SA, SWG

Visualization: DC, HJS, BJ, NPM, DS, EBK, EK, VSFA, AB, SWG, LHT

Funding acquisition: LHT Project administration: DS

Supervision: LHT, DC

Writing – original draft: DC, HJS, BJ, NPM, DS, EBK, EK, VSFA

Writing – review & editing: DC, HJS, BJ, NPM, DS, EBK, EK, VSFA, BD, LHT

## Data and materials availability

All data are available in the main text or the supplementary materials.

## METHODS

### Study Design

We report results from initial safety, feasibility and optimization studies to evaluate induced entrainment in response to 40 Hz sensory stimulation performed at the Massachusetts Institute of Technology (MIT) (NCT04042922) and the University of Iowa and a longitudinal study designed to evaluate the effects of long-term 40 Hz stimulation in participants with mild AD (NCT04055376), as described in the demographics below and in the Supplemental Materials in full. Both studies were registered on clinicaltrials.gov (NCT04042922; NCT04055376). All studies were approved by the Committee on the Use of Humans as Experimental Participants (COUHES) at the MIT and the University of Iowa Institutional Review Board (IRB), respectively. These studies were carried out in accordance with the Code of Ethics of the World Medical Association (Declaration of Helsinki). All participants and their primary caregivers provided written informed consent before participation.

For the initial safety, feasibility and optimization study performed at MIT (NCT04042922), each participant had surface electroencephalogram (EEG) recordings while receiving sensory stimulation from our device (see EEG section below). Sensory stimulation was delivered using a system composed of a white light emitting diode (LED) panel (2’x2’, natural white color, 3900-4000K) that was modified for flicker and brightness control (Neltner Labs) and a sound system (SB2920-C6, VIZIO, or LP-2024A+, Lepy connected to BRS40, BOSS).

The longitudinal study registered on clinicaltrials.gov was a single-blinded, randomized, placebo-controlled study in which participants were given stimulation devices used 1-hour daily at home over the course of 9 months (NCT04055376). The placebo group used devices delivering constant light and white noise while the active group used devices delivering synchronized combined light and sound stimulation at 40 Hz, both set at the same light and sound levels. The primary outcome measures for this study was safety and compliance of usage. Safety was assessed with adverse events questionnaires completed over weekly phone calls with staff and evaluations for aberrant ictal spikes in response to stimulation using electroencephalogram (EEG) at baseline and 3 months. Compliance was assessed using built-in time-stamp log of when the device is on or off. Exploratory outcomes included effects on cognition, structural changes and functional connectivity in the brain, and sleep. Participants underwent baseline assessments including a neuropsychiatric battery, EEG, magnetic resonance imaging (MRI) and blood collection for sequencing. Participants wore actigraphy devices throughout the study. Safety was assessed with weekly calls. The neuropsychiatric battery, EEG and MRI was repeated at month 3 however, due to the COVID-19 pandemic, EEG and MRI could not be performed after the 3-month timepoint. Here, we report results from the 3-month timepoint.

### Participants

This study involved three cohorts of participants. The first cohort included three groups of volunteers who were recruited at the Massachusetts Institute of Technology (MIT): (1) cognitively normal adults between the ages of 18-35 (young group); (2) cognitively normal adults between the ages of 50-100 (older group); and (3) adults with a previous diagnosis of probable AD, between the ages of 50-100, with a Mini-Mental State Examination (MMSE) (Folstein, Folstein, and McHugh 1975) score of 19-26 at screening (mild AD group). Main exclusion criteria were: (1) active treatment with N-methyl-D-aspartate (NMDA) receptor antagonist (e.g., memantine), anti-epileptic agent, or psychiatric agent (e.g., antidepressant, antipsychotic); and (2) history of seizure or stroke within 24 months prior to the study participation (full criteria listed in Table S1). Participants in the mild AD group were additionally assessed using the Montreal Cognitive Assessment (MoCA) (Nasreddine et al. 2005) and the clinical dementia rating scale (CDR) (Hughes et al. 1982). Participants in this cohort completed a brief cognitive assessment and then were recorded using scalp electroencephalogram (EEG) during single-(visual or auditory) and multi-sensory (synchronized visual and auditory) stimulation at 40 Hz.

The second cohort included neurosurgical patients with medically intractable epilepsy who were recruited at the University of Iowa Hospitals & Clinics. These patients were being admitted to the hospital for 7-14 days for invasive monitoring with intracranial electrodes to localize their seizure focus. An intracranial EEG monitoring plan was generated by the University of Iowa Comprehensive Epilepsy Program. Following intracranial electrode implantation surgery, patients remained in an electrically-shielded epilepsy monitoring room in the University of Iowa’s Clinical Research Unit. After the final surgical treatment plan was agreed upon between the clinical team and the patient, 1-2 days before the planned resection and after the patient had restarted anti-epileptic medications, the patient was recorded using intracranial EEG during single- and multi-sensory stimulation at 40 Hz.

The third cohort included patients who met the National Institute of Aging-Alzheimer’s Association and Diagnostic and Statistical Manual of Mental Disorders, Fifth Edition (DSM-5) clinical criteria for probable mild AD dementia were recruited at MIT. The diagnosis was confirmed by the investigator based on interviews with the patient and their caregiver along with neurological assessments as part of the study. These patients were between the ages of 50-100 and had the MMSE score between 19-26 at screening. Main exclusion criteria were: (1) MMSE score outside the range of 19-26 at screening; (2) history of seizure; and (3) refusal to participate in the study protocol at screening (full criteria listed in Table S1).

Participants enrolled in the longitudinal study were patients with probable mild AD dementia with the same criteria described above.

### Evaluation of induced entrainment

#### Scalp EEG Recording with sensory stimulation

Participants underwent EEG recording while receiving sensory stimulation that was delivered using a system composed of a white light emitting diode (LED) panel (2’x2’, natural white color, 3900-4000K) that was modified for flicker and brightness control (Neltner Labs) and a sound system (SB2920-C6, VIZIO, or LP-2024A+, Lepy connected to BRS40, BOSS). The LED panel and the speaker were controlled by a custom circuit board (Neltner Labs) that housed Teensy USB Development Board (version 3.6, PJRC). Participants were seated five feet from the device during the EEG recording. Each participant was exposed to three 40 Hz stimulation conditions (visual, auditory, and combined) and three non-40 Hz conditions (visual, auditory, and combined), with each condition lasting for about 3 minutes (for cognitively normal participants) or 1 minute (for patients with mild AD). For the combined stimulation conditions, auditory and visual pulses were temporally aligned at their onset.

Scalp EEG was recorded throughout the stimulation experiments using an ActiveTwo system (Biosemi), with 32 scalp electrodes arranged according to the international 10-20 system. Electrooculogram (EOG) electrodes were used to monitor eye blinks and lateral eye movements, with one EOG electrode placed near the infraorbital ridge of the left eye and another electrode placed near the lateral canthus of the right eye. Two additional electrodes were placed over the mastoids bilaterally. The magnitude of the offset value was kept below 40 mV for each electrode. EEG signals were recorded with a low-pass hardware filter with a half-power cutoff at 104 Hz and then digitized at 512 Hz with 24 bits of resolution. The data were saved to a computer along with the “trigger events” that were manually inserted during the experiment to denote the start and finish of each stimulation period.

#### Intracranial EEG Recording with sensory stimulation

At the University of Iowa, patients with medically intractable epilepsy underwent an anatomical and functional MRI scanning session within two weeks of surgery with a GE discovery MR750W 3 Tesla scanner. After the final surgical treatment plan was agreed upon between the clinical team and the patient, typically 1-2 days before the planned resection and after the patient had restarted anti-epileptic medications, participants underwent sensory stimulation with our device. Each participant was exposed to three 40 Hz stimulation conditions (visual, auditory, and combined), with each condition lasting for about 1 minute. Intracranial EEG was recorded throughout the stimulation experiments using electrode arrays (Ad-Tech Medical Instrument) that included stereotactically-implanted depth electrodes (4-8 macro contacts per electrode, spaced 5-10 mm apart) and grid arrays (containing platinum-iridium disc contacts, with 2.3 mm exposed diameter, 5-10 mm inter-contact distance, embedded in a silicon membrane) placed on the cortical surface. A subgaleal electrode was used as a reference. Data acquisition was controlled by an ATLAS Neurophysiology system (NeuraLynx). Collected data were amplified, filtered (0.1-500 Hz bandpass), digitized at 2 kHz, and stored for subsequent offline analysis.

#### Detection of Epileptiform Discharges

All EEG recordings were reviewed for the presence of epileptiform discharges by an expert neurophysiologist blinded to the timing and the duration of the stimulation. For scalp EEG recordings, raw signals without any preprocessing were examined for epileptiform discharges. For intracranial EEG, due to strong line noise at 60 Hz, raw signals were bandstop filtered between 59 and 61 Hz and bandpass filtered between 0.1 and 100 Hz before examination.

#### MRI

For the Phase 2 study, MRI data were acquired using a 3-Tesla Siemens Tim Trio scanner (Siemens, Erlangen, Germany) paired with a 12-channel phased-array whole-head coil. Head motion was restrained with foam pillows. Subjects who did not undergo an MRI scan at both baseline and 3-month visit were excluded from this analysis for a total number of 13 subjects (Active n = 7; control n = 6). Sequences included 3D T1-weighted magnetization prepared rapid acquisition gradient echo (MP-RAGE) anatomical images, Functional T2-weighted images were acquired using a gradient-echo echo-planar pulse sequence sensitive to bold oxygenation level-dependent (BOLD) contrast (Kwong et al. 1992; Ogawa et al. 1992) were acquired. To allow for T1-equilibration effects, 4 dummy volumes were discarded prior to acquisition. Functional resting data were acquired while the participant was instructed to rest with eyes open for a period of 5 minutes consisting of 120 volumes. Functional task data was recorded during the visual presentation of face-name stimuli described below for a total of 98 volumes per run. Online prospective acquisition correction (PACE) was applied to the EPI sequence. Structural MRI data was analyzed using FSL and Freesurfer. Ventricular enlargement analysis was conducted using the VIENA pipeline within FSL version 5.0.9. Functional MRI data was analyzed using CONNToolbox using a seed-driven approach (Whitfield-Gabrieli and Nieto-Castanon 2012) and Statistical Parametric Mapping (SPM - https://www.fil.ion.ucl.ac.uk/spm/). Images were realigned (motion corrected), spatially normalized to the Montreal Neurological Institute (MNI) stereotactic space, and smoothed with a five mm kernel. Quality assurance was performed on the functional time series in order to detect outliers in the motion and global signal intensity using Conn. Additionally, subjects were removed from analysis if the number of valid scans made up less than 10% of the total recorded scans. Resulting in a total number of 12 subjects for the resting state analysis (Active n = 6, control n = 6).

#### Actigraphy

Sleep and Activity were assessed using an actigraphy device (ActiGraph Link GT9X+, Firmware 1.7.2, ActiGraph, Pensacola, FL) worn on the non-dominant wrist and worn continuously throughout the study. Five, 7-day recordings: the first 7-days after home installation of the stimulation device (baseline), 30 days and 60 days later (Month 1, Month 2), a median of 101 days, range: 85-109 (Month 3) aligned following their visit and 120 (±15 days) (Month 4) were used for analysis. We confirmed participants used the stimulation panel in at least 6 of the 7 days in which we analyzed their activity data, as referenced by their device log. Data were visually inspected to confirm availability and wear during these periods followed by Wear Time Validation preprocessing (ActiLife software version 6.13.4, ActiGraph, Pensacola, FL) using 90 minute non-wear threshold (Choi et al. 2012; Knaier et al. 2019). Participants who had wear time below 60% of total recording length (n = 1) or an incomplete recording for any of the timepoint data collections (n = 3) were excluded from analysis. Data collected beginning in April 2020 and later was excluded because of the anticipated effects of the COVID-19 pandemic among our patient population.

Non-parametric analyses of actigraphy were used because of the statistical characteristics of activity counts (Van Someren et al. 2019; Gonçalves et al. 2014). Our analysis used Y-axis acceleration data from the actigraph, sleep/wake epochs using the Cole-Kripke algorithm (ActiLife) and compiled code from National Sleep Research Resource, (actiCircadian) implemented in MATLAB R2020a (Mathworks, Natick, MA).

Individual days within a subject’s recording are excluded from this analysis if there were fewer than 10 hours of wear during daytime or more than 1 hour of non-wear in the major sleep period (9 pm to 7 am) as determined by the program. Interdaily stability (IS) was calculated as the ratio between the variance of the average 24-h pattern and the overall variance, a measure of circadian rhythmicity, higher values indicate more stability. Interdaily variability (IV) was calculated as the ratio between the mean square first derivative and the overall variance; it and quantifies fragmentation of rhythms, a lower value indicates fewer disruptions during the day.

#### Face-Name Association Task and Delayed Recall

The functional task consisted of viewing faces unfamiliar to study participants paired with fictional first names in a modified novel vs. repeated design as described in Sperling et al. (Sperling et al. 2001). Each face-name pair was presented for 5 seconds followed by a 0.8 second period of fixation to a white, central crosshair. Participants viewed seven different face-name pairs per novel or repeated blocks. The repeated block consisted of alternation between two face-name pairs (one male, one female), while the novel condition consisted of new face-name pairs for each stimuli presentation across all runs. Up to six runs of the block design paradigm exhibited below were conducted for each participant. Participants were given explicit instructions to try to remember the name associated with each face.

Immediately following the imaging session, participants were tested on a subset of the face-name pairs. 14 pairs from the novel face-name sets presented were shown with two names listed below. Participants were asked to choose the name that had been presented with the face during the imaging session. A subset of participants was not able to complete the task due to the duration of the MRI session or missing scan during 3-month visit and were removed from this analysis. The total number of participants used in this analysis was 9 (Active n = 4, control n =5)

#### Neuropsychological testing

A comprehensive neuropsychological test battery was administered by the study neurologist at the initial visit (M0). Assessments included the Mini Mental State Exam (MMSE; (Folstein, Folstein, and McHugh 1975)), Montreal Cognitive Assessment (MoCA; (Nasreddine et al. 2005)) and the following measures from the National Alzheimer’s Coordination Center’s Uniform Data Set version 3.0 (Weintraub et al. 2018): Trail Making Test Part A (TMT-A, time in seconds); TMT Part B (TMT-B, time in seconds); Craft 21 Story Recall: Immediate (Story Recall Immediate, 44-item); Craft 21 Story Recall: Delayed (Story Recall Delayed, 44-item); Number Span Test Forwards (NST-F, longest span); Number Span Test Backwards (NST-B, longest span); Geriatric Depression Scale (GDS, 15-item); Functional Assessment Scale (FAS, 30-item); Neuropsychiatric Inventory Questionnaire (NPI-Q); and Clinical Dementia Rating (CDR). The Alzheimer’s Disease Assessment Scale-cognitive subscale (ADAS-Cog Total, 70 item) was also administered and sub-scores were calculated for word list immediate recall (ADAS-Cog Recall, 10-item) and delayed recognition (ADAS-Cog Recognition, 12-item; (Graham et al. 2004) A briefer assessment battery consisting of the MMSE, MoCA, ADAS-Cog, Number Span Test, TMT (A and B), CDR, NPI-Q, and GDS was completed at month 3 (median of 101 days, range: 85-109).

#### Sequencing

Genomic DNA was purified from approximately 5×10^6 peripheral white blood cells using a genomic DNA extraction kit (Qiagen, 69506). Extracted DNA was then submitted to Genewiz for SNP genotyping at rs429358. 14 subjects were genotyped. (n=1) subject was excluded from analysis due to an insufficient blood draw.

#### Statistical Analysis

Group-level PSD and coherence results were expressed as the median across participants within each group. 95% confidence intervals for the median were obtained by bootstrapping across the participants 60,000 times with replacement. For comparisons between stimulation conditions or participant groups, if a condition or group was involved in more than one family-wise comparisons, the Friedman test (for paired comparisons) or the Kruskal-Wallis test (for unpaired comparisons), followed by Dunn’s multiple comparison test was used to calculate the p values. Otherwise, the Wilcoxon’s sign rank test (for paired comparisons) or the Mann-Whitney test (for unpaired comparisons) was used. For comparisons at multiple electrode sites or electrodes, the Bonferroni correction was applied by multiplying the p values by the number of electrode sites or electrodes. P values for categorical variables were calculated using Fisher’s exact test.

For MRI analyses, unpaired t-tests were used for group comparisons of ventricular volume while paired t-tests were used for group level analyses of hippocampal volume and mean functional connectivity to seed regions using resting-state fMRI. Functional connectivity data was FWE-corrected and p < 0.05 was considered significant.

For actigraphy, we performed a 2-way ANOVA (Treatment Group x Time) with Sidak’s multiple comparisons to assess between or within group differences over time. Delta comparisons are the value at a given timepoint minus the Baseline value for any variable of interest. Comparison of this value to 0, or no change was used to assess if the stimulation protocol had a significant impact within each group.

A two-sided p value less than 0.05 was considered significant. Specific statistical tests and parameters are detailed in the figure legends. All statistical analyses were performed using MATLAB 2019b (MathWorks, Inc.) and GraphPad Prism 8.4 (GraphPad Software).

