## Supplemental Data and Methods for "Gamma Frequency Sensory Stimulation in Probable Mild Alzheimer’s Dementia Patients: Results of a Preliminary Clinical Trial" (from another version of this preprint: version 2, 2021-05-11)

### SUPPLEMENTAL FIGURES

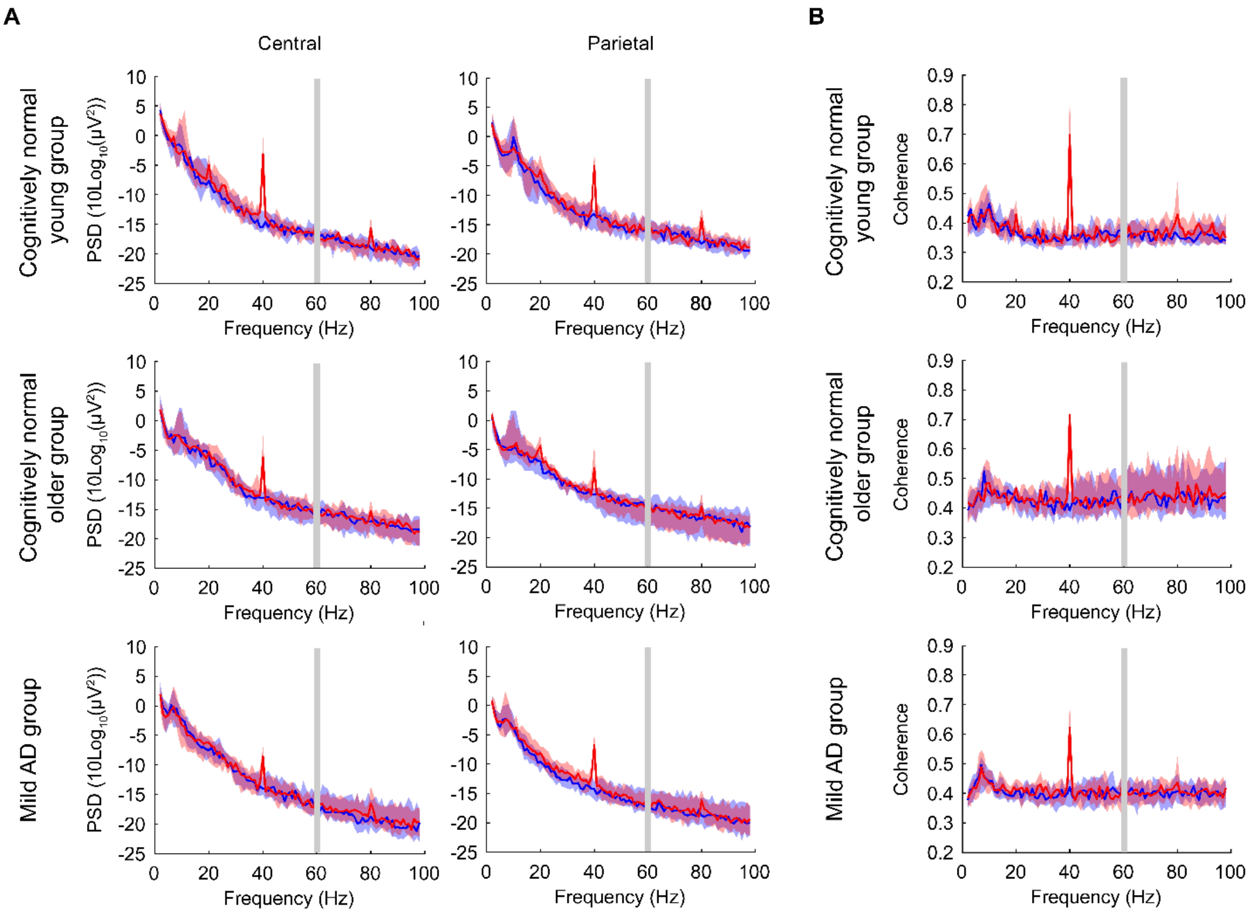

**Figure S1. Changes in scalp EEG power spectral density and coherence with acute 40 Hz combined stimulation, related to Figure 1.**

(A) Scalp EEG power spectral density (PSD) at the central (Cz, C3, C4) and parietal (Pz, P3, P4, P7, P8) electrode sites, in cognitively normal young subjects (n = 13; top row), cognitively normal older subjects (n = 12; middle row), and patients with mild AD (n = 16; bottom row). Solid lines, group median; shaded areas, 95% confidence interval; blue, no stimulation; red, 40 Hz combined stimulation. Gray bar placed around frequency range with 60 Hz line noise.

(B) Scalp EEG global coherence in cognitively normal young subjects (n = 13; top row), cognitively normal older subjects (n = 12; middle row), and patients with mild AD (n = 16; bottom row). Solid lines, group median; shaded areas, 95% confidence interval; blue, no stimulation; red, 40 Hz combined stimulation. Gray bar placed around frequency range with 60 Hz line noise.

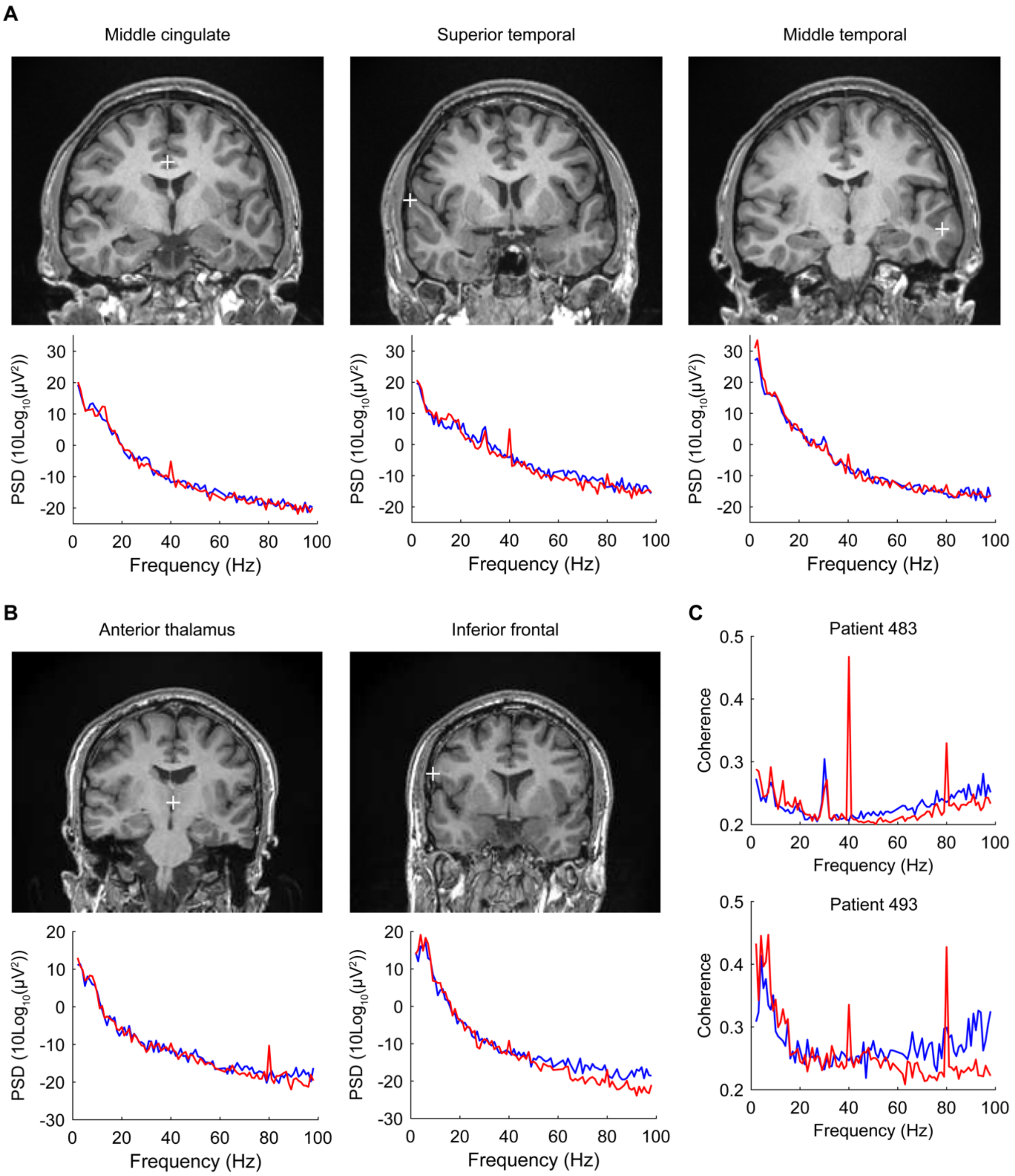

**Figure S2. Intracranial EEG power spectral density and coherence during no stimulation and acute 40 Hz combined stimulation, related to Figure 1.**

(A, B) Example coronal MRI images before electrode implantation (top row) and intracranial EEG power spectral density (PSD; bottom row) from Patient 483 (A) and Patient 493 (B), for depth electrode contacts placed in deep and superficial brain regions. White plus sign, approximate location of the depth electrode contact; blue, no stimulation; red, 40 Hz combined stimulation. For the PSD between 58 Hz and 62 Hz, interpolated

values are plotted, because bandstop filtering around 60 Hz during signal preprocessing (for line noise removal) led to a large dip in the PSD around 60 Hz.
(C) Global coherence in Patient 483 (left) and Patient 493 (right). Blue, no stimulation; red, 40 Hz combined stimulation. For the global coherence between 58 Hz and 62 Hz, interpolated values are plotted, because bandstop filtering around 60 Hz during signal preprocessing (for line noise removal) led to a large dip in the global coherence around 60 Hz. Change in global coherence at 40 Hz from the no-stimulation level was 0.26 for Patient 483 and 0.09 for Patient 493.
(D) Heat maps showing the change in 40 Hz coherence with 40 Hz combined stimulation compared to no stimulation for all electrode contact pairs, in Patient 483 (left) and Patient 493 (right). Number of contact pairs showing an increase in 40 Hz coherence from the no-stimulation level was 7807 (out of 9045 possible pairs) for Patient 483 and 237 (out of 351 possible pairs) for Patient 493.

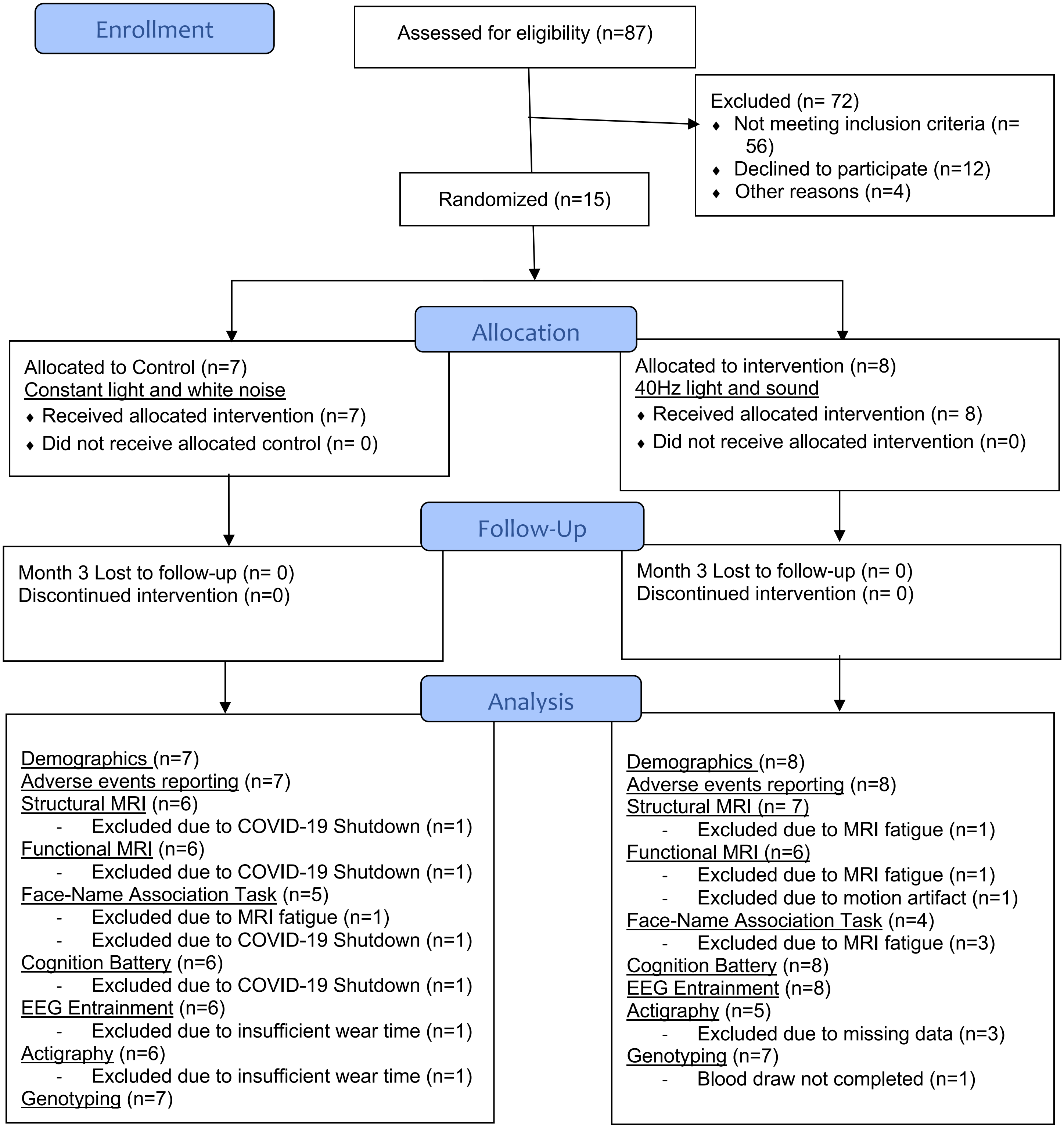

**Figure S3. CONSORT Flow Diagram: Longitudinal Cohort**
Participant recruitment, allocation, follow-up and analysis counts by activity

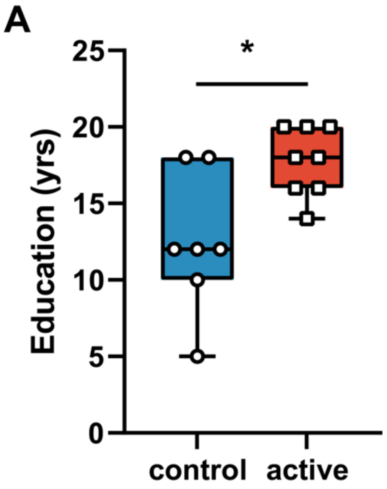

**Figure S4. Participant's years of education**

Results of an unpaired t-test, $*p < 0.05$. $p = 0.012$ ($n = 15$, control = 7, active = 8).

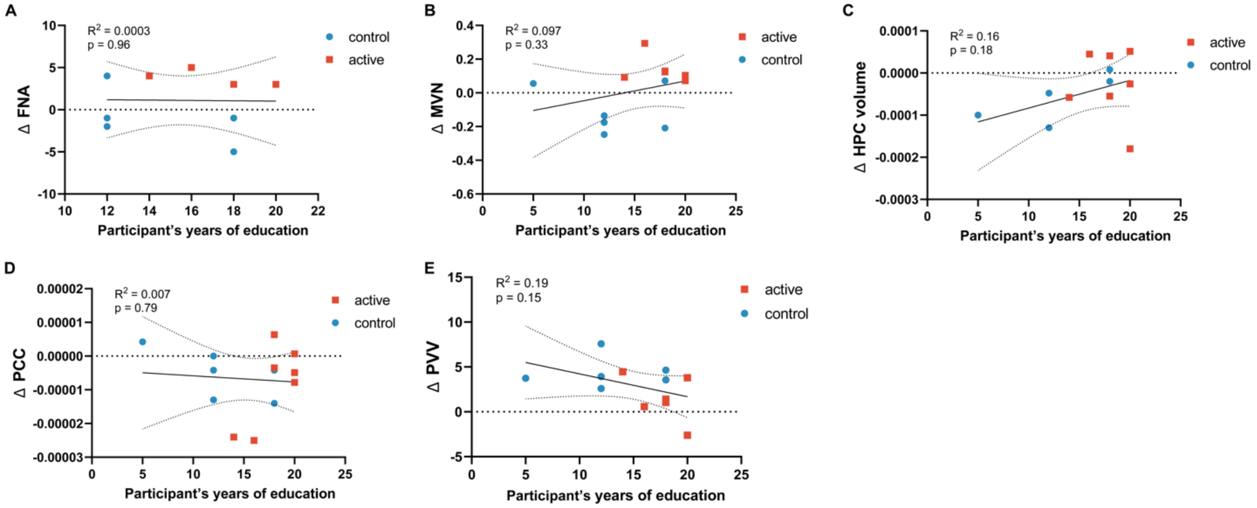

**Figure S5. Years of education related to Face-Name Association and MRI outcomes**

(A) Simple linear regression of participant's level of education (years) vs change in Face-Name Association. Goodness of Fit $R^2$ = 0.0003, p = 0.96. Solid black line represents best fit line, dotted black represents 95% Confidence Interval.

(B) Simple linear regression of participant's level of education (years) vs change in Medial Visual Network Connectivity. Goodness of Fit $R^2$ = 0.097, p = 0.33. Solid black line represents best fit line, dotted black represents 95% Confidence Interval.

(C) Simple linear regression of participant's level of education (years) vs change in Bilateral Hippocampal Volume. Goodness of Fit $R^2$ = 0.16, p = 0.18. Solid black line represents best fit line, dotted black represents 95% Confidence Interval.

(D) Simple linear regression of participant's level of education (years) vs change in Posterior Cingulate Cortex Connectivity. Goodness of Fit $R^2$ = 0.007, p = 0.79. Solid black line represents best fit line, dotted black represents 95% Confidence Interval.

(E) Simple linear regression of participant's level of education (years) vs change in Ventricular Volume. Goodness of Fit $R^2$ = 0.19, p = 0.15. Solid black line represents best fit line, dotted black represents 95% Confidence Interval.

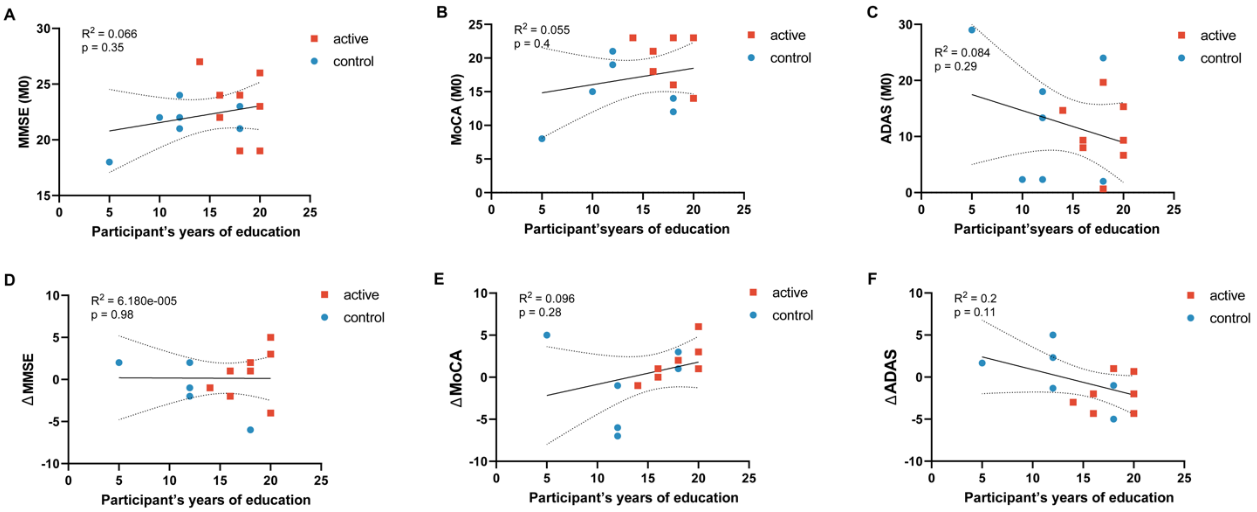

**Figure S6. Years of education related to cognition at baseline and change in cognition at Month 3 Visit**

(A) Simple linear regression of participant's level of education (years) vs baseline Mini-Mental Status Exam. Goodness of Fit $R^2$ = 0.066, p = 0.35. Solid black line represents best fit line, dotted black represents 95% Confidence Interval.

(B) Simple linear regression of participant's level of education (years) vs baseline Montreal Cognitive Assessment. Goodness of Fit $R^2$ = 0.0055, p = 0.4. Solid black line represents best fit line, dotted black represents 95% Confidence Interval.

(C) Simple linear regression of participant's level of education (years) vs baseline Alzheimer's Disease Assessment Scale. Goodness of Fit $R^2$ = 0.084, p = 0.29. Solid black line represents best fit line, dotted black represents 95% Confidence Interval.

(D) Simple linear regression of participant's level of education (years) vs change in MMSE. Goodness of Fit $R^2$ = 6.180e-005, p = 0.98. Solid black line represents best fit line, dotted black represents 95% Confidence Interval.

(E) Simple linear regression of participant's level of education (years) vs change in MOCA. Goodness of Fit $R^2$ = 0.096, p = 0.28. Solid black line represents best fit line, dotted black represents 95% Confidence Interval.

(F) Simple linear regression of participant's level of education (years) vs change in ADAS. Goodness of Fit $R^2$ = 0.2, p = 0.11. Solid black line represents best fit line, dotted black represents 95% Confidence Interval.

A

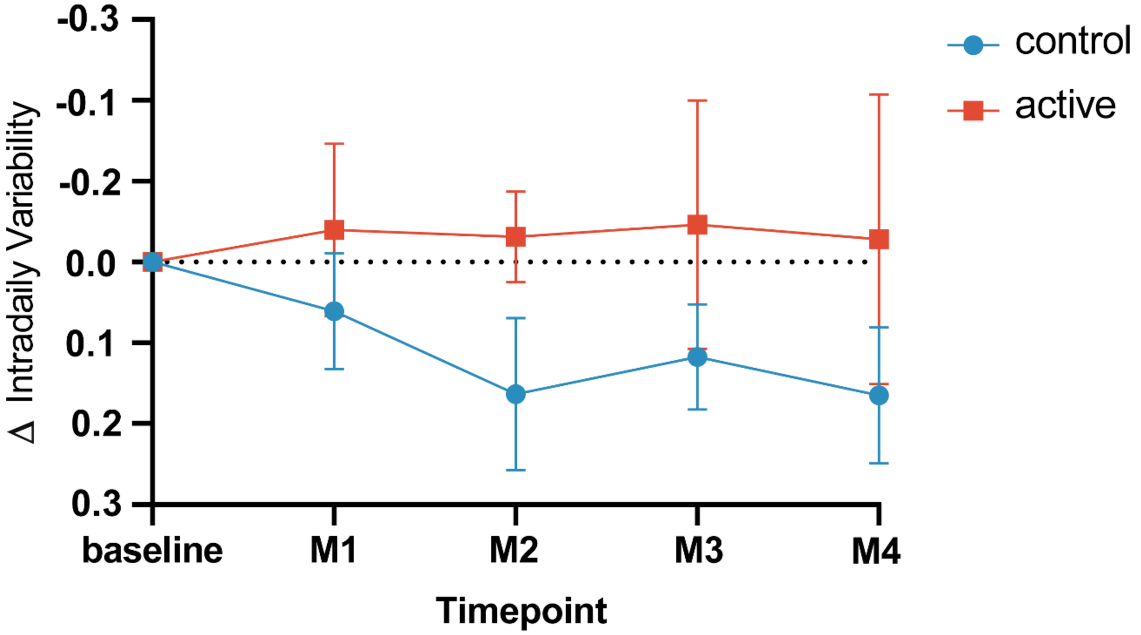

**Figure S7. Actigraphy Data for Circadian Rhythm Fragmentation.**
Change in Intradaily Variability demonstrates a trend towards stabilization in the active group but no change in control group. Statistical analysis with a 2-Way ANOVA with Šídák's multiple comparisons at each timepoint compared to 0 (no-change). Lines indicate means ± SEM. Mean of control = 0.13, mean of active = -0.04. Main effect of intervention, p = 0.22, interaction of intervention x time, p = 0.88, M4 multiple comparison, control, p = 0.30, active, p = 0.99.

| Study group | Inclusion criteria | Exclusion criteria |
|---|---|---|
| **Cognitively normal young and older groups** | • Subject is between the ages of 18 - 35 and 50 - 100<br>• Subject is willing and able to sign informed consent form | • Subjects who are pregnant.<br>• Subjects who do not have healthcare.<br>• Subjects who are being treated with acetylcholinesterase inhibitors. (donepezil, galantamine, rivastigmine) or N-methyl-D-aspartate (NMDA) receptor antagonists (eg. Memantine) who have been on a stable dose for less than 60 days.<br>• Subjects with history of seizure or epilepsy within the past 24 months.<br>• Active treatment with one or more anti-epileptic agent.<br>• Subjects who have had a stroke within the past 24 months.<br>• Subjects diagnosed with a known diagnosis of migraine headache.<br>• Active treatment with one or more psychiatric agent (e.g. antidepressants, antipsychotics, etc).<br>• Subjects who have an active implantable medical device including but not limited to implantable cardioverter defibrillator (ICD), deep brain stimulator (DBS), cardiac pacemaker, and/or sacral nerve stimulator.<br>• Subjects who have contraindications for MRI imaging.<br>• Subjects who have a life expectancy of less than 2 years. |
| **Mild AD group** | • Subject is between the ages of 50 - 100.<br>• Subject must have mild Alzheimer's disease with a Mini Mental State Exam (MMSE) score of 19 -26.<br>• Subject is willing to sign informed consent document.<br>• If subject is deemed to not have capacity to sign the informed consent, he/she will need a legally authorized representative to provide surrogate consent. | • Subjects who do not have healthcare.<br>• Subjects who are being treated with N-methyl-D-aspartate (NMDA) receptor antagonists (eg. Memantine).<br>• Subjects on medications that lower seizure threshold such as wellbutrin, ciprofloxacin, levofloxacin, etc.<br>• Subjects with history of seizure or epilepsy within the past 24 months. |

| | | |
|---|---|---|
| | | • Subjects with clinically significant suicide risk and/or suicide attempt in the past 1 year.<br>• Subjects with behavioral problems such as aggression/agitation/impulsivity that might interfere with their ability to comply with protocol.<br>• Active treatment with one or more anti-epileptic agent.<br>• Subjects who have had a stroke within the past 24 months.<br>• Subjects diagnosed with migraine headache.<br>• Active treatment with one or more psychiatric agent (e.g. antidepressants, antipsychotics, etc).<br>• Subjects who have an active implantable medical device including but not limited to implantable cardioverter defibrillator (ICD), deep brain stimulator (DBS), cardiac pacemaker, and/or sacral nerve stimulator.<br>• Subjects who have profound hearing or visual impairment.<br>• Subjects who have a life expectancy of less than 2 years.<br>• Subjects who are pregnant. |

**Table S1. Eligibility criteria.**

| | Change in 40 Hz Global Coherence from No Stimulation, Median (Range)[a] | 40 Hz Combined Stimulation vs No Stimulation p Value[b] |
|---|---|---|
| **Young, CN (n = 13)** | 0.34 (0.17 to 0.61) | <0.001 |
| **Older, CN (n = 12)** | 0.23 (0.09 to 0.41) | <0.001 |
| **Mild AD (n = 16)** | 0.22 (0.04 to 0.45) | <0.001 |

Abbreviations: CN, cognitively normal; AD, Alzheimer's disease.
[a] Ranges from -1 to 1, with a more positive value indicating more increase in the 40 Hz global coherence and a more negative value indicating more decrease in the 40 Hz global coherence with 40 Hz stimulation compared to no stimulation.
[b] Wilcoxon's sign rank test.

**Table S2. Changes in scalp EEG global coherence at 40 Hz with acute 40 Hz combined stimulation, related to Figure 1.**

Resting-State Functional Connectivity Changes

| Region | MNI coordinates | | | #Voxels | Peak T | Pval (size) |
|---|---|---|---|---|---|---|
| | X | Y | Z | | | |
| PCC - Ctrl - Ses1>Ses2 | | | | | | |
| rSMG post. | 52 | -40 | 44 | 79 | -3.04 | 0.000 |
| rFG mid. | 46 | 32 | 24 | 46 | -2.5 | 0.000 |
| lAG | -40 | -54 | 42 | 31 | -2.53 | 0.000 |
| lLOC sup. | -40 | -60 | 56 | 22 | -0.75 | 0.003 |
| rFG mid. | 46 | 26 | 32 | 17 | -2.65 | 0.020 |
| rFG sup. | 4 | 26 | 46 | 16 | -2.25 | 0.029 |
| lFP | -40 | 34 | 8 | 15 | -3.75 | 0042 |
| PCC - Active - Ses1>Ses2 | | | | | | |
| N/A | | | | | | |
| PCC - Active (Ses1>Ses2) > Ctrl (Ses1>Ses2) | | | | | | |
| lFP | -42 | 34 | 8 | 36 | 7.32 | 0.031 |
| rFP | 50 | 40 | 4 | 35 | 7.01 | 0.032 |
| Left HPC – Ctrl – Ses1>Ses2 | | | | | | |
| N/A | | | | | | |
| Left HPC – Active – Ses1>Ses2 | | | | | | |
| lLOC sup. | -20 | -86 | 24 | 27 | 12.64 | 0.001 |
| rLOC sup. | 28 | -86 | 32 | 16 | 17.37 | 0.006 |
| Left HPC – Active (Ses1>Ses2) > Ctrl (Ses1>Ses2) | | | | | | |
| N/A | | | | | | |
| Bil. HPC – Ctrl – Ses1>Ses2 | | | | | | |
| N/A | | | | | | |
| Bil. HPC – Active – Ses1>Ses2 | | | | | | |

| | | | | | | |
|---|---|---|---|---|---|---|
| lLOC sup. | -20 | -84 | 22 | 62 | 7.51 | 0.000 |
| lOP | -32 | -92 | 2 | 32 | 6.06 | 0.016 |
| Bil. – Active (Ses1>Ses2) > Ctrl (Ses1>Ses2) | | | | | | |
| N/A | | | | | | |

**Table S3. Resting-State Functional connectivity clusters.**
From top to bottom indicating regions showing change in functional connectivity with the posterior hub of the default mode network including the posterior cingulate cortex (PCC) and precuneus, change in control group connectivity between sessions, change in active group connectivity between sessions, and change between groups between sessions. Followed by functional connectivity changes for left and bilateral hippocampus. Right hippocampus showed no changes across all comparisons. $P < 0.05$ FWE-corrected for all clusters, paired T-test.
SMG: Supramarginal Gyrus
FG: Frontal Gyrus
AG: Angular Gyrus
LOC: Lateral Occipital Cortex
FP: Frontal Pole
OP: Occipital Pole

| | Control | Active | P-value |
|---|---|---|---|
| Left HPC | 1.84e-3(+/-3.27e-4) | 1.64e-3(+/-1.85e-4) | 0.204 |
| Right HPC | 2.00e-3(+/-4.02e-4) | 1.67e-3(+/-2.63e-4) | 0.099 |
| Bil. HPC | 3.84e-3(+/-7.14e-4) | 3.31e-3(+/-4.32e-4) | 0.127 |
| Med. Vis. FC | 0.09(+/-0.09) | 0.06(+/-0.06) | 0.588 |
| FNA recall | 7.80(+/-1.48) | 6.25(+/-1.89) | 0.209 |
| Left HPC | 1.84e-3(+/-3.27e-4) | 1.64e-3(+/-1.85e-4) | 0.204 |
| Right HPC | 2.00e-3(+/-4.02e-4) | 1.67e-3(+/-2.63e-4) | 0.099 |
| Bil. HPC | 3.84e-3(+/-7.14e-4) | 3.31e-3(+/-4.32e-4) | 0.127 |
| Med. Vis. FC | 0.09(+/-0.09) | 0.06(+/-0.06) | 0.588 |
| FNA recall | 7.80(+/-1.48) | 6.25(+/-1.89) | 0.209 |

**Table S4. Baseline comparisons between groups for MRI related outcomes.**
Data are represented as mean +/- standard deviation. p-values generated using t-test
HPC: Hippocampus
Med. Vis.: Medial Visual Network
FC: Functional Connectivity
FNA: Face-name association

| | Change in Hippocampal Volume (Percent) | | |
|---|---|---|---|
| | Median (Range) | 25th Percentile | 75th Percentile |
| Control (n=6) | -1.94 (4.09) | -2.81 | -0.34 |
| Active (n=7) | -0.87 (6.52) | -1.97 | 1.41 |
| | Between group comparison p-value: 0.034 | | |
| | Percent Ventricular Volume Change | | |
| | Median (Range) | 25th Percentile | 75th Percentile |
| Control (n=6) | 3.84(4.98) | 3.31 | 5.37 |
| Active (n=7) | 1.23(7.08) | -0.226 | 3.96 |
| | Between group comparison p-value: 0.024 | | |

**Table S5. Structural MRI Statistics**
Data are represented as median (range)
p-values generated using paired t-test for hippocampal volume and unpaired t-test for ventricular volume
